# Development and validation of decision rules models to stratify coronary artery disease, diabetes, and hypertension risk in preventive care: cohort study of returning UK Biobank participants

**DOI:** 10.1101/2021.03.01.21252657

**Authors:** José Castela Forte, Pytrik Folkertsma, Rahul Gannamani, Sridhar Kumaraswamy, Sarah Mount, Tom J. de Koning, Sipko van Dam, Bruce H. R. Wolffenbuttel

**Affiliations:** Department of Clinical Pharmacy and Pharmacology, University of Groningen, University Medical Center Groningen, the Netherlands; Ancora Health B.V., Groningen, The Netherlands; Department of Endocrinology, University of Groningen, University Medical Center Groningen, the Netherlands; Department of Neurology, University of Groningen, University Medical Center Groningen, the Netherlands; Pediatrics, Department of Clinical Sciences, Lund University, Lund, Sweden

**Author notes:** **Correspondence:** José Castela Forte, BSc (Hon), Department of Pharmacy and Pharmacology, Hanzeplein 1, P.O. Box 30.001, 9700 RB Groningen, The Netherlands.

**Keywords:** Coronary artery disease, Hypertension, Diabetes, Prevention medicine, Risk stratification

## Abstract

**Background:** A wide range of predictive models exist that predict risk of common lifestyle conditions. However, these have not focused on identifying pre-clinical higher risk groups that would benefit from lifestyle interventions and do not include genetic risk scores.

**Objective:** To develop, validate, and compare the performance of three decision rule algorithms including biomarkers, physical measurements and genetic risk scores for incident coronary artery disease (CAD), diabetes (T2D), and hypertension in the general population against commonly used clinical risk scoring tools.

**Methods:** We identified 60782 individuals in the UK Biobank study with available follow-up data. Three decision rules models were developed and tested for an association with incident disease. Hazard ratios (with 95% confidence interval) for incident CAD, T2D, and hypertension were calculated from survival models. Model performance in discriminating between higher risk individuals suitable for lifestyle intervention and individuals at low risk was assessed using the area under the receiver operating characteristic curve (AUROC).

**Results:** We ascertained 500 incident CAD cases, 1005 incident T2D cases, and 2379 incident cases of hypertension. The higher risk group in the decision rules model had a 40-, 40.9-, and 21.6-fold increase in risk of CAD, T2D, and hypertension, respectively (*P* < 0.001 for all). Risk increased significantly between the three strata for all three conditions (*P* < 0.05). Risk stratification based on decision rules identified both a low-risk group (only 1.3% incident disease across all models), as well as a high-risk group where at least 72% of those developing disease within 8 years would have been recommended lifestyle intervention. Based on genetic risk alone, we identified not only a high-risk group, but also a group at elevated risk for all health conditions.

**Conclusion:** We found that decision rule models comprising blood biomarkers, physical measurements, and polygenic risk scores are superior at identifying individuals likely to benefit from lifestyle intervention for three of the most common lifestyle-related chronic health conditions compared to commonly used clinical risk scores. Their utility as part of digital data or digital therapeutics platforms to support the implementation of lifestyle interventions in preventive and primary care should be further validated.

## Introduction

Developed countries have seen a consistent rise in life expectancy and overall improving trends in chronic disease outcomes [1]. In just six decades, this has translated to a global increase in life expectancy of over 20 years for both men and women [1]. Yet, longer life expectancy has been accompanied by an increase in the prevalence of common chronic diseases, such as coronary artery disease (CAD), type 2 diabetes (T2D), and hypertension, which pose a significant burden to societies and limit healthy life expectancy (HALE) [2,3]. Preventive strategies which allow for earlier lifestyle intervention are a solution to tackle the growing burden of lifestyle-related health conditions. Indeed, lifestyle interventions such as weight loss, limiting (saturated) fat intake, and 30 minutes of exercise per day are recommended across multiple guidelines to reduce cardiovascular disease risk and the progression from prediabetes to T2D [4,5]. Yet, the sustainable implementation of lifestyle interventions faces several challenges, and cannot be achieved with one-size-fits-all approaches [6]. Rather, adherence and maintenance of health behaviour change requires personalized lifestyle recommendations.

To be able to provide such targeted lifestyle recommendations, the first step is to adequately stratify risk in individuals in a pre-clinical state and prioritize which aspects of their health they ought to focus on. For the three prevalent chronic health conditions mentioned above, several risk assessment tools have been made available to primary care physicians, including the Framingham risk scores [7,8,9]. These risk scores incorporate clinical and laboratory parameters, and have been shown to perform comparably well in European populations to other risk scores [7,10]. However, over two thirds of models for cardiovascular risk are restricted to a mixture of demographics, medical history, blood pressure and lipid profile, and a limited set of lifestyle factors, such as smoking [11]. Until now, these models do not include physical measurements or genetic susceptibility, although these health conditions are known to be multifactorial in nature, and for instance, progression from prediabetes to T2D is accelerated by even modest increases in adiposity, in individuals at higher genetic risk [6]. Especially when several studies have shown that the addition of genetic risk scores, as well as scores combining physical measurements and lifestyle factors, to demographic and biomarker data can improve risk stratification in preventive and primary care settings [11,12,13,14,15,16,17].

This study aimed to evaluate decision rules models incorporating other routine biomarkers, physical measurements, and genetic information in addition to established risk factors and investigate whether these improve risk stratification for three prevalent lifestyle-related health conditions in the large population-based UK Biobank cohort.

## Methods

### Study population

The UK Biobank is a longitudinal population-based cohort of 502,503 participants aged between 37–73 years old, collected between 2006 and 2010. For this study, we included only participants without coronary artery disease, T2D, and hypertension diagnosed by a physician at recruitment, in whom extensive follow-up data were available. In addition, individuals without diagnosed disease but who at baseline crossed a “clinical threshold” for any of the health conditions were also excluded from further analysis. These were individuals with a systolic blood pressure between 140 and 180 mmHg systolic or between 90 and 120 mmHg diastolic for hypertension [18], a fasting glucose value above 7.0 mmol/L for T2D [19], individuals with significantly impaired kidney function for cardiovascular disease [20,21]. Individuals for whom any of the variables in Table 1 were missing were also excluded. This study was conducted under UK Biobank application 55495, and followed TRIPOD reporting guidelines (Appendix S1). Local Institutional Review Board ethics approval was not necessary for this study.

**Table 1.**
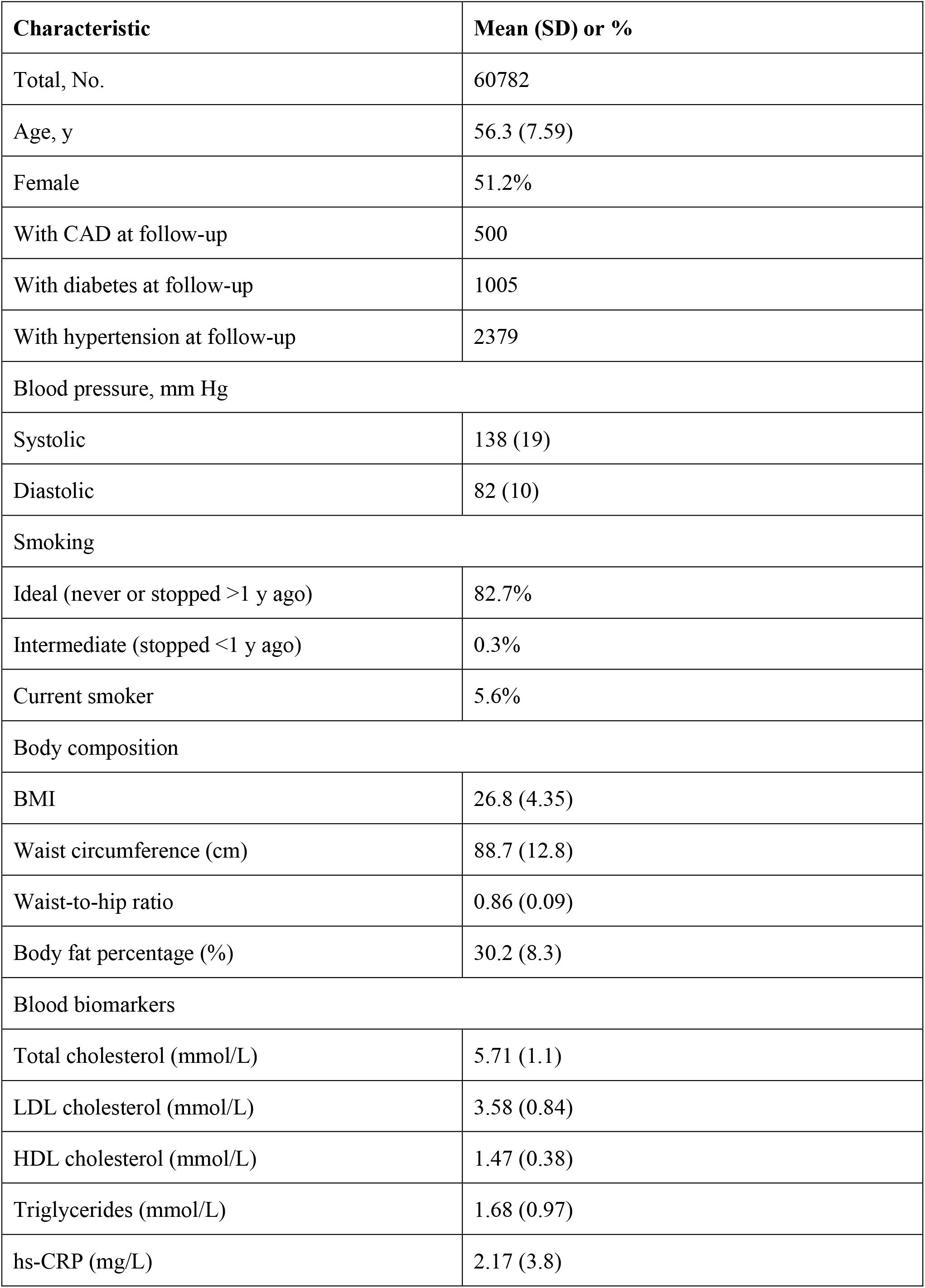
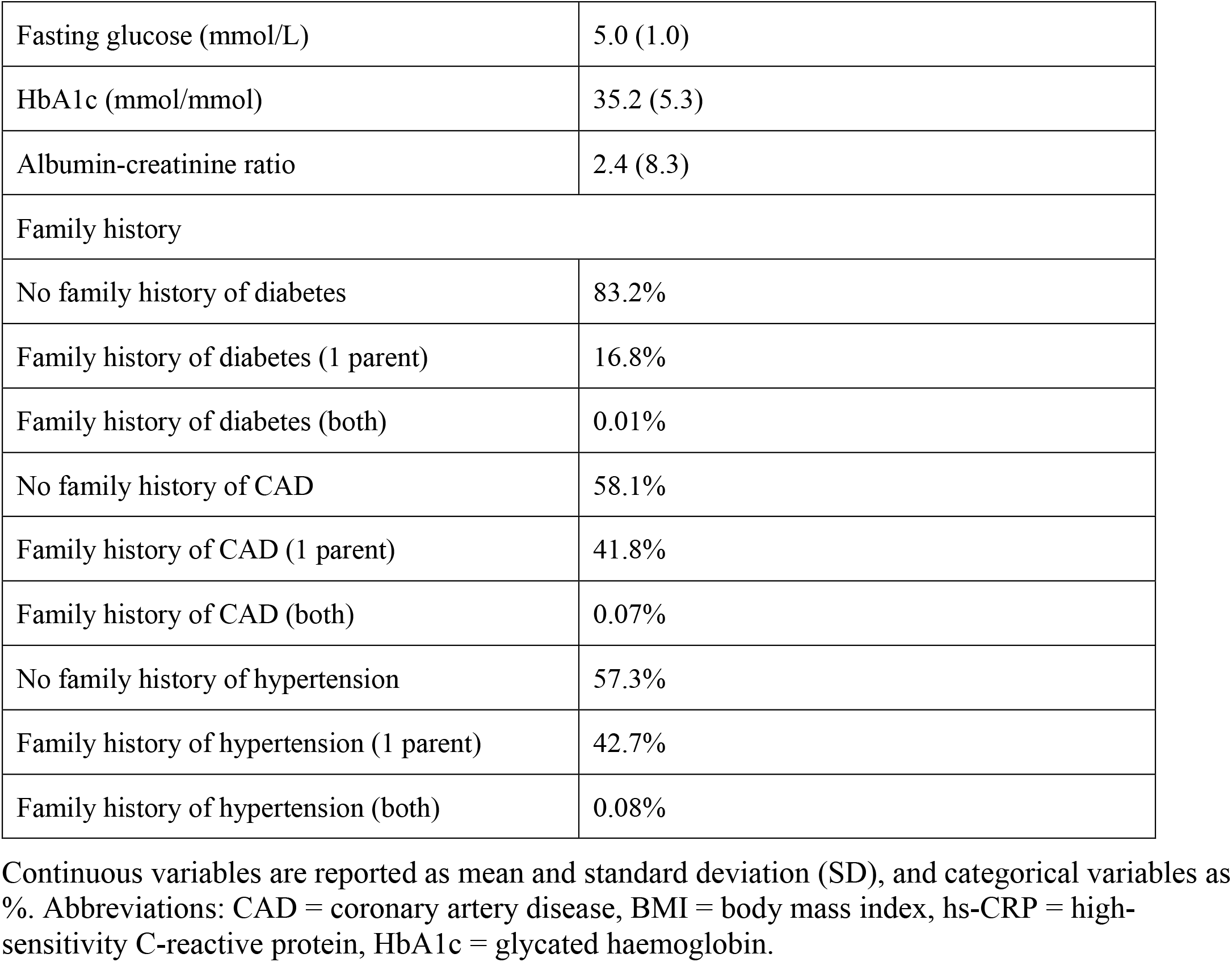
Baseline characteristics table.

| **Characteristic** | **Mean (SD) or %** |
| --- | --- |
| Total, No. | 60782 |
| Age, y | 56.3 (7.59) |
| Female | 51.2% |
| With CAD at follow-up | 500 |
| With diabetes at follow-up | 1005 |
| With hypertension at follow-up | 2379 |
| Blood pressure, mm Hg |  |
| Systolic | 138 (19) |
| Diastolic | 82 (10) |
| Smoking |  |
| Ideal (never or stopped >1 y ago) | 82.7% |
| Intermediate (stopped <1 y ago) | 0.3% |
| Current smoker | 5.6% |
| Body composition |  |
| BMI | 26.8 (4.35) |
| Waist circumference (cm) | 88.7 (12.8) |
| Waist-to-hip ratio | 0.86 (0.09) |
| Body fat percentage (%) | 30.2 (8.3) |
| Blood biomarkers |  |
| Total cholesterol (mmol/L) | 5.71 (1.1) |
| LDL cholesterol (mmol/L) | 3.58 (0.84) |
| HDL cholesterol (mmol/L) | 1.47 (0.38) |
| Triglycerides (mmol/L) | 1.68 (0.97) |
| hs-CRP (mg/L) | 2.17 (3.8) |
| Risk stratification for cardiovascular disease |  |
| Fasting glucose (mmol/L) | 5.0 (1.0) |
| HbA1c (mmol/mol) | 35.2 (5.3) |
| Albumin-creatinine ratio | 2.4 (8.3) |
| Family history |  |
| No family history of diabetes | 83.2% |
| Family history of diabetes (1 parent) | 16.8% |
| Family history of diabetes (both) | 0.01% |
| No family history of CAD | 58.1% |
| Family history of CAD (1 parent) | 41.8% |
| Family history of CAD (both) | 0.07% |
| No family history of hypertension | 57.3% |
| Family history of hypertension (1 parent) | 42.7% |
| Family history of hypertension (both) | 0.08% |

Continuous variables are reported as mean and standard deviation (SD), and categorical variables as %. Abbreviations: CAD = coronary artery disease, BMI = body mass index, hs-CRP = high-sensitivity C-reactive protein, HbA1c = glycated haemoglobin.

### Biomarkers, physical measurements and polygenic risk scores

To define the risk factors for each of the health conditions, a literature search was conducted in accordance with the 2009 Preferred Reporting Items for Systematic Reviews and Meta-Analyses (PRISMA) statement [21]. We searched for meta-analyses indexed in PubMed that were published between January 2014 and end of 2019 (additional details on the search strategy available in table S1, and PRISMA flowcharts in Figs S1-3). We also searched relevant national and international clinical guidelines not originally identified by the search. Based on the findings of the literature review, rules were defined to stratify individuals as high, elevated, and no elevated risk. These rules as described in detail in Appendix S2, and shortly below. Data on biomarkers was retrieved from the blood biochemistry category in the UKB, physical measurements from the body size measurements and abdominal composition categories, and smoking status was ascertained based on the self-reported smoking status registered at recruitment. Family and medical history were retrieved from the respective categories.

#### Coronary artery disease

For coronary artery disease, the literature and additional guideline search identified total cholesterol [22], HDL cholesterol [7,23,24], LDL cholesterol [24], triglycerides [23,25,26,27], and high-sensitivity C-reactive protein (hs-CRP) [28,29,30] as relevant blood biomarkers. The Framingham Risk Score for 10-year coronary heart disease risk was used, which included information on treatment for hypertension and smoking status [7]. A polygenic risk score for coronary artery disease was calculated as described below. Individuals were classified as high risk for which intervention is advised if they met any of the following rules, all weighted equally: total cholesterol above 8 mmol/L, systolic blood pressure above 180 mmHg, LDL cholesterol above 4.9 mmol/L, the incidence risk according to Framingham was high and the genetic susceptibility score being was “high”, or if biomarkers other than LDL were out of range. The no elevated risk profile was defined as no biomarkers being out of range, the genetic susceptibility score below the eighth decile, and negative family history. All others for which at least one risk factor was elevated were classified as intermediate risk.

#### Diabetes

Glycaemic variables (fasting glucose and HbA1c), blood lipids, markers of body composition, blood pressure, family history, gender, and smoking were identified as risk factors [8,31,32,33,34,35]. The Framingham Risk Score for diabetes was used, and a polygenic risk score for diabetes was calculated [8]. Participants were placed in the highest stratum if they met any of the following rules, all weighted equally: HbA1c was above 6.5% and fasting glucose was below 6.1mmol/L, fasting glucose was above 6.1mmol/L, either of the glycaemic variables was elevated (HbA1c between 5.5% and 6.4% or fasting glucose between 5.6 mmol/L and 6.1mmol/L) and they were overweight/obese, their clinical risk was high, their glucose was unregulated and their genetic susceptibility was high, or if their clinical risk was elevated, they were older than 45, had a HDL cholesterol below 0.9 mmol/L, and triglycerides above 2.8 mmol/L [19]. Participants were classified as not being at elevated risk if all biomarkers were within normal range, the genetic susceptibility score was below the eighth decile, and clinical risk was not elevated. All others with at least one marker out of range were considered at intermediate risk.

#### Hypertension

For hypertension, the literature and additional guideline search identified age [36], systolic and diastolic blood pressure [36,37,38,39,40], body mass index (BMI) [41,42], gender [43], and smoking status as relevant markers. The Framingham Risk Score for hypertension risk [9] was used, and a PRS for systolic blood pressure was calculated. Participants were classified as high risk if their systolic blood pressure was between 130-140 mmHg, or the diastolic between 80 and 90 mmHg. Equally, those with a high clinical risk, or an elevated clinical risk and a high PRS were stratified as high risk. The no elevated risk profile was defined as all biomarkers being within normal range, the genetic susceptibility score being below the eighth decile, and incidence risk according to the clinical score not being elevated. All others with at least one marker out of range were considered at intermediate risk.

### Polygenic risk scores

Polygenic risk scores (PRS) were calculated using an additive model for CAD, T2D and hypertension. Individuals were binned into deciles based on their PRS scores and the average disease incidence was calculated for each decile. The difference between individuals in the tenth risk decile, those in the nineth and eighth deciles, and all other deciles were assessed. The 1000 Genomes dataset was used as reference panel for the LD calculations [44]. The genotyping data and data containing the tested phenotype outcomes were downloaded from the UKB. All variants with an imputation R^2^ < 0.4 determined with the minimac3 algorithm, were removed from the downloaded genotyping files [45]. Summary statistics files from three large genome-wide association studies (GWAS) conducted in other cohorts were used to calculate PRS for CAD, T2D, and hypertension [46,47,48]. These publicly available summary statistics were reformatted where necessary to be consistent with the format required by LDpred. A rho of 1 was used, and all variants with a GWAS significance p-value below 0.01 were selected based on previous studies showing marginal differences between this and other stringency cutoffs (table S4) [13]. In total, the T2D, CAD, and hypertension PRS included 199120, 139885 and 400016 SNPs, respectively. The PRS were also computed with and without adjustment of the following variables: genotyping array, first four principal components, age and sex. To assess the added predictive value of PRS over sex and age alone, we also added the predictions of a logistic regression model including only sex and age (table S2). Individuals were binned into deciles based on their PRS scores and the average disease incidence at each age was calculated for each decile (Fig S1). Additional methods are available in Appendix S3.

### Ascertainment of disease incidence

Information regarding the variables used to calculate incidence for each of the health conditions at 8 years after study enrollment can be found in table S2. In short, we used International Statistical Classification of Diseases and Related Health Problems (ICD) codes, and the self-reported diagnoses collected at recruitment and follow-up questionnaires.

### Statistical analysis

Similar to the three strata of risk for the decision rules model, three risk strata (“low”, “intermediate” and “high”) were defined for the Framingham scores. For the coronary artery disease risk score, the bottom, middle, and top tertiles were used as risk categories [7]. For diabetes, categories were based on <3%, 3% to 8%, and >8% incidence risk at eight-years [8]. For hypertension, this was <5%, 5% to 10%, and >10% incidence risk [9].

To evaluate the ability to discriminate higher risk individuals who would be suggested lifestyle intervention from those at no elevated risk, we used the area under the receiver operating characteristics (AUROC) curve computed from 2000 bootstrap iterations. Sensitivity and specificity for each model is also presented. Cox proportional hazards models were used to test the association of risk strata defined by the decision rules model and the clinical scores with incident events of CAD, T2D, and hypertension. Hazard ratios (HRs) with 95% confidence intervals were calculated between risk strata and the reference group (those not at elevated risk for the decision rules model, and low risk for Framingham).

We considered a p-value <0.01 as statistically significant for differences in AUROC determined by DeLong’s nonparametric test and p-value <0.05 significant for differences in risk between strata [49]. All data analyses were performed using R software v4.0.3 and the “*survival”, “survminer” and* “ggplot2” packages were used to conduct the survival analysis and generate graphs.

## Results

### Population characteristics

In total, 60 782 unique participants had follow-up data available, of which 42 978, 36 913, and 33 541 were included in the analyses for T2D, CAD, and hypertension, respectively. Table 1 shows the baseline characteristics of the study population. At follow-up, there were 500 incident CAD cases, 1005 incident T2D cases, and 2379 incident hypertension cases. Participants were aged 56.3 years on average, and slightly more participants were female (51.2%). Average values for all lipid markers were above general recommendations [21]. Similarly, physical measurements of BMI and waist circumference were above the existing thresholds for abdominal obesity, and both average systolic and diastolic blood pressure values crossed the stage 1 hypertension threshold [50,51].

### Polygenic risk scores

For all three health conditions, a higher PRS was strongly associated with a higher incidence rate (Fig 1). For the highest risk stratum compared to the rest of the population, this translated to a hazard ratio (HR) of 4.6 (95% CI 3.8-5.6), 2.9 (2.5-3.4), and 1.9 (1.7-2.1) for CAD, T2D, and hypertension, respectively (table 2). When comparing the highest risk individuals to those in the first seven deciles, the HRs were 7 (95% CI 5.7-8.7), 3.8 (3.2-4.4), and 2.2 (2.0-2.5) (table 3). The risk for individuals in the ninth and eighth deciles was also significantly higher compared to individuals in the first seven deciles, with HRs of 3.4 (2.7-4.2) for CAD, 2.3 (2.0-2.7) for T2D, and 1.8 (1.7-2) for hypertension (table 3).

**Table 2.** Risk increase for the individuals at high genetic risk (10th decile), compared to individuals at low genetic risk (1-7th decile) of population. Second and third column show hazard ratios calculated based on a logistic regression model adjusted for the respective variables. In all cases the difference with the remainder of the population was statistically significant (p-value < E-100).

|  | **Unadjusted PRS** | **PRS adjusted for 4 PCs and array type** | **PRS adjusted for 4 PCs, array type, sex and age** | **Age and sex** |
| --- | --- | --- | --- | --- |
| **CAD** | 1.66 (0.93-2.35) | 2.25 (1.39-3.11) | 4.43 (3.14-5.74) | 2.55 (1.63-3.47) |
| **T2D** | 1.86 (1.33-2.39) | 2.61 (1.96-3.26) | 2.81 (2.12-3.50) | 1.47 (1.00-1.94) |
| **Hypertension** | 1.37 (1.06-1.62) | 1.61 (1.30-1.92) | 1.77 (1.45-2.09) | 1.50 (1.21-1.79) |

CAD: coronary artery disease, T2D: Type 2 diabetes, PRS: polygenic risk scores, PC: genetic principal component.

**Table 3.** Area under the receiver operating characteristic (AUROC) curve for model discrimination of higher risk individuals amenable to lifestyle intervention assessed for the clinical risk score(s), PRS, and decision rules model. Number of individuals classified as low and higher risk and number of individuals who had developed disease at follow-up are also presented.

| Model and health condition | N low risk individuals | N low risk who developed disease | N recommend lifestyle intervention | N recommended lifestyle intervention who developed disease | AUROC (95% CI) |
| --- | --- | --- | --- | --- | --- |
| **CAD (N = 21969 women, 167 cases; 14944 men, 333 cases)** |  |  |  |  |  |
| FRS women | 7426 | 22 | 5680 | 99 | 0.67 (0.63-0.71) |
| FRS men | 5171 | 55 | 4431 | 165 | 0.60 (0.58-0.63) |
| PRS | 25839 | 173 | 3692 | 165 | 0.62 (0.60-0.64) |
| Rule model | 1521 | 0<sup>s</sup> | 14980 | 360 | 0.66 (0.64-0.68) |
| **Diabetes (N = 42978, 1005 cases)** |  |  |  |  |  |
| FRS | 12305 | 39 | 12634 | 726 | 0.72 (0.71-0.73) |
| PRS | 30084 | 467 | 4298 | 239 | 0.57 (0.56-0.58) |
| Rule model | 8351 | 13 | 14169 | 819 | 0.75 (0.74-0.76) |
| **Hypertension (N = 33541, 2379 cases)** |  |  |  |  |  |
| FRS | 3359 | 23 | 26587 | 2317 | 0.60 (0.59-0.60) |
| PRS | 23479 | 1327 | 3354 | 391 | 0.54 (0.53-0.54) |
| Rule model | 2274 | 17 | 12506 | 1759 | 0.70 (0.69-0.71) |

*Abbreviations: N = number; FRS = Framingham risk score; PRS = polygenic risk score. AUROC reported with 95% confidence interval*
*\* indicates statistically significant difference in performance between decision rules model and PRS,*
*# indicates statistically significant difference in performance between decision rules model and clinical risk score(s), <sup>s</sup>computed as 1 for statistical analysis purposes.*

**Figure 1.**
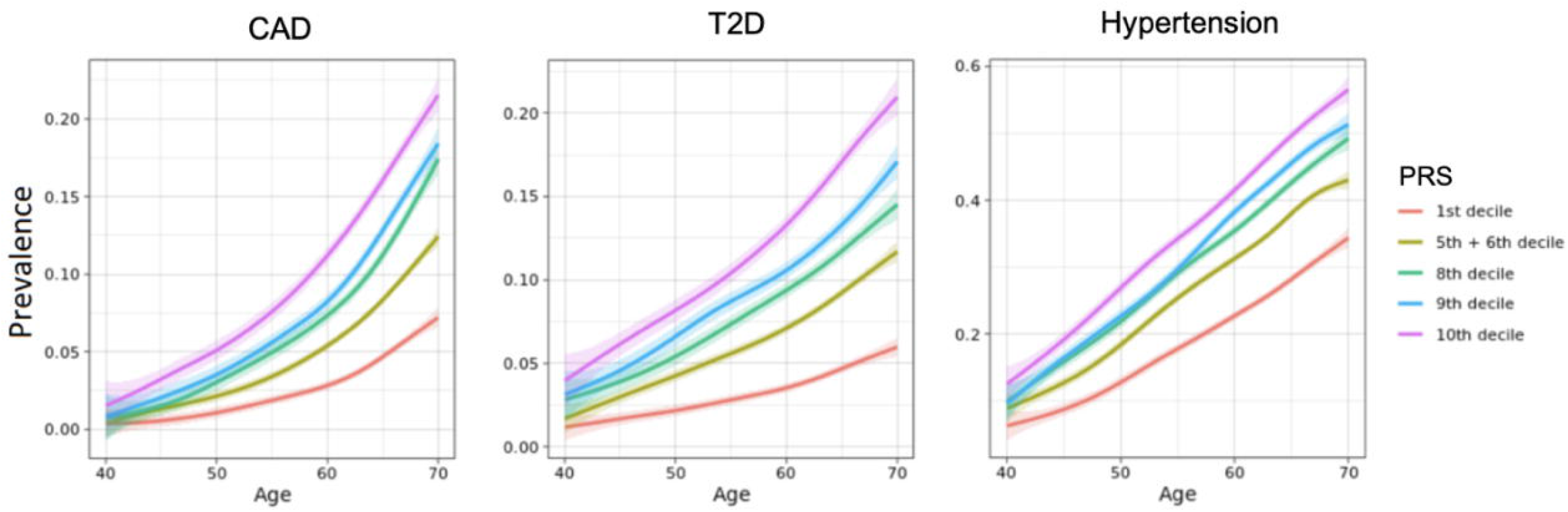
Cumulative incidence in different risk strata. Risk classification conducted based on the logistic regression model of the PRS adjusted for age, sex, first four principal components and array type.

### Sensitivity analysis

The Framingham scores achieved an AUROC of 0.67 (95% CI 0.63-0.71) for women and 0.60 (0.58-0.63) for men, 0.72 (0.71-0.73), and 0.60 (0.59-0.60) for CAD, T2D, and hypertension, respectively. Sensitivity and specificity for these models were 59.3% and 74.4%, 49.5% and 70.8%, 72.2% and 71.6%, and 97.4% and 22.1%. The performance of the decision rules model was better than Framingham for CAD in men, T2D, and hypertension with an AUROC of 0.66 (0.64-0.68) for CAD, 0.75 (0.74-0.76) for T2D, and 0.70 (0.69-0.71) for hypertension (table 2, P < 0.01 for all). There was no difference in performance between the decision rules model and Framingham for CAD in women. The discriminatory power of the decision rules model was also superior to PRS alone (table 3). Specificity was higher for T2D (68.2%, 67.7%-68.6%) and hypertension (65.5%, 65%-66%), with sensitivity also higher for T2D (81.5%, 79.1%-84%) and lower for hypertension (73.9%, 72.7%-75.7%). For CAD, the decision rules model achieved higher sensitivity (72.0%, 68.0%-76.0%) but lower specificity (59.8%, 59.3%-60.3%) than both Framingham models. For the decision rules models, positive predictive values were higher for T2D, hypertension and CAD in women, but lower than for the men’s model (table 4). Negative predictive values were extremely high for all models, with the highest being 99.58% (99.50%-99.65%) for the Framingham for CAD in women and the lowest 97.05% (96.85%-97.24%) for the decision rules for hypertension (table 4).

### Risk stratification and lifestyle advice recommendations

The observed absolute risk for each health condition differed between the high, intermediate, and low risk strata for the decision rules model, but not for the clinical risk score (Fig 2). In terms of absolute risk, being classified as high risk by the clinical score translated to a 2.6% and 1.4% difference in absolute risk compared to not being at elevated risk for CAD in men (HR 3.8, 2.8-5.1) and in women (HR 6.8, 4.3-10.8), 2.1% for T2D (HR 3.7, 2.1-6.7), and 7.4% for hypertension (HR 14.1, 9.36-21.3). For the intermediate risk stratum, there was a risk difference for CAD in men and women, but not for T2D or hypertension. In comparison, the high-risk group in the decision rules model showed a 2.34% increase in absolute risk for CAD (HR 40, 5.6-283), 5.64% for T2D (HR 40.9, 23.7-70.8), and 12.4% for hypertension (HR 21.6, 13.4-34.8). For the intermediate risk group, these differences were 0.62% (HR 4, 1.5-77), 0.69% (HR 5.6, 3.2-9.9), and 2.4% (HR 4.5, 2.8-7.3), respectively. If all individuals in the higher risk group were recommended lifestyle intervention as a consequence of their baseline measurements, 40.6%, 33%, and 37.2% of all individuals would be recommended lifestyle intervention for CAD, T2D and hypertension with the decision rules model. For T2D and hypertension, this is 41.6% and 53% less than if the clinical risk scores were used, while detecting as many cases for T2D and only 561 fewer for hypertension. For CAD, 14980 (40.6%) of individuals would have been recommended intervention by the decision rules, compared to 10111 (27.4%) for Framingham. This represents a difference of detecting and advising intervention to 72% of all those who eventually developed disease, as opposed to 53.2%. In addition, 15.4% of those who ended up developing CAD were classified as low risk by Framingham, compared to 0.2% for the decision rules model.

**Figure 2.**
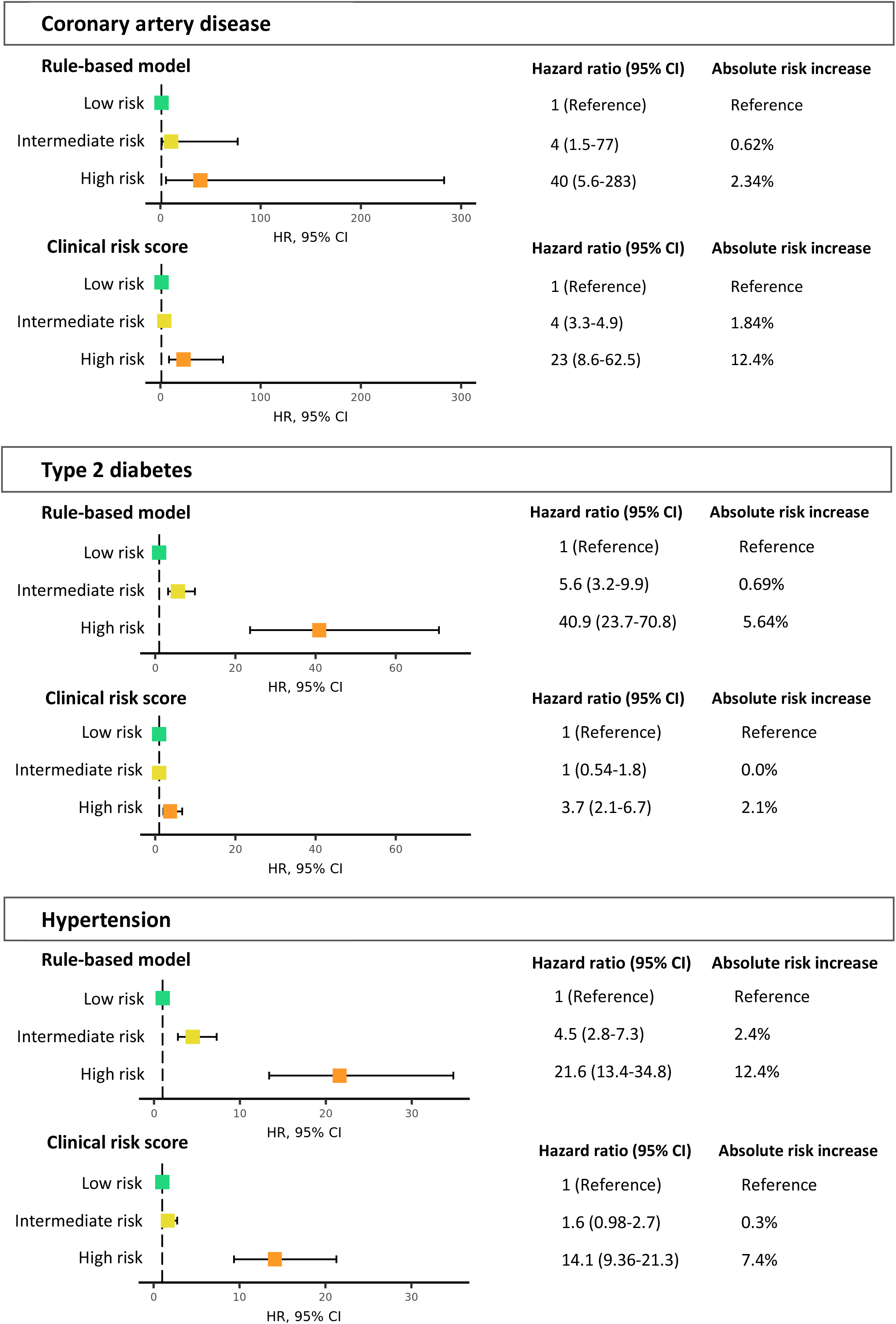
Hazard ratios (HR) of disease incidence per risk stratum. The group with no risk factors is used as reference. Both the HR and the absolute risk are displayed for each decision rules model and clinical score for all three health conditions. CI = confidence interval.

## Discussion

We investigated the association of different risk categories of three decision rule models incorporating blood biomarkers, physical measurements, and genetic information, with incident disease for three common lifestyle-related health conditions and compared its performance to currently used clinical risk calculators in 60782 returning participants in the population-based UK Biobank study. Individuals classified as high risk who would be recommended lifestyle intervention by the decision rules model had a 40-, 41- and 22-fold higher 8-year risk of CAD, T2D, and hypertension compared to those who were classified as not having elevated risk. All decision rules models either outperformed the respective Framingham clinical score or showed improvement in the detection of cases likely to benefit from lifestyle intervention.

We showed that adding other biomarkers, physical measurements, and genetic risk to traditional clinical risk scores leads to moderate improvement in predictive performance. From the many clinical risk scores for risk estimation of cardiometabolic health conditions available, we chose to compare our decision rules models to the Framingham risk scores due to their extensive validation across multiple cohorts [10,11]. In this sub-population of the UKB cohort, the Framingham scores performed comparably to studies in North American and Dutch populations (0.63 to 0.67), but slightly worse than reports from other studies, including lower than in the best original validation studies (0.66 to 0.83) [58,59]. For hypertension, specifically, the inferior performance of the Framingham model compared to other studies likely comes from the substantially lower number of prehypertensive individuals and mean blood pressure values in these cohorts (below 120 mmHg systolic and 75 mmHg diastolic) compared to the UKB, leading to as many as 79.3% of individuals being classified as high risk [56,57]. Compared to the Framingham scores, all three decision rules models improved either the detection of cases likely to benefit from lifestyle intervention or of those least likely to do so [52,53]. The slight improvement in performance of the rules model for diabetes is not surprising, as unregulated HbA1c is a risk factor for disease development in prediabetics, and specific insulin resistance phenotypes are linked to central adiposity. Similarly, there is growing evidence for genetics playing a more central role in the diabetes burden than previously thought [54,55]. For hypertension, the addition of genetic data also likely explains the improved performance of the rules model.

While modest in magnitude, the differences in performance between the different models could have significant practical implications. Preventive health programs should consider the health risks of individuals holistically across a spectrum of mental and physical health. By improving the precision to detect those who eventually developed the disease and would be recommended intervention and minimizing the number of individuals who did not develop the disease and would have nonetheless been advised to take action, these models have the potential to increase the impact of such programs in two ways. On the one hand, it could increase their effectiveness, since the number of prevented cases if the interventions were successfully implemented would be higher. On the other hand, cardiometabolic health issues are highly prevalent. By also accurately identifying individuals less likely to benefit from a cardiometabolic health intervention in the short-term, these models can be combined with models for other physical and mental health conditions and help low risk individuals to prioritize lifestyle changes in other aspects of their health. With recent studies showing that programs as short as three to five months can trigger diabetes remission and improve cardiovascular risk factors [60,61,62], the use of these stratification mechanisms for a periodic risk assessment across varied lifestyle conditions would be a valuable tool for optimizing return on investment in personalized preventive medicine programs.

With regards to the addition of genetic risk to clinical scores, our findings support recent studies that suggested adding genetic susceptibility scores to clinical scores for CAD and T2D, as well as stroke or cardiovascular disease led to improvements in risk prediction [12,63,64]. Based on genetic risk alone, we identified a group of high risk individuals with hazard ratios of 4.6-, 2.9-, and 1.9 for CAD, T2D and hypertension. However, we also encountered differences between the top risk decile and the ninth and eighth deciles, and between these and the rest of the population. In comparison, Khera et al. identified a similar risk increase only in the top 8% and 3.5% of individuals in the UKB, for CAD and T2D respectively, and the top 5% individuals in the Finnish cohort of Mars et al. were at 2.62-fold increased risk of CAD and 3.28-fold for T2D (table S3, Fig S3) [12,13]. This effectively demarcates not only a “high risk”, but also an “elevated risk” group in these two deciles, compared to the “no elevated risk” group comprising the rest of the population.

One of the barriers to the implementation of risk models in preventive and primary care has been the belief that such algorithms have an actual low impact on decision-making in apparently healthy individuals, and mostly generate demand for “unnecessary” care [65,66,67]. In this study, we make two significant contributions to help overcome this issue. First, we showed that easily interpretable decision rules models including genetic risk can better identify individuals at low risk unlikely to benefit from lifestyle interventions in the short-term than traditional clinical scores. Models based on risk factor burden are easy to interpret and communicate, and a simple metric such as the absence or presence of more than one risk factor is associated with substantial differences in lifetime risk of cardiometabolic health conditions [68]. By including genetic risk in risk factor burden calculations in an additive way, we can identify individuals at genetically elevated or high risk with normal demographic and blood risk factors. There remain substantial financial and technical challenges in conducting GWAS, and in correctly calculating and interpretating PRS, for different health conditions. In individuals for whom routinely collected medical and biomarker data clearly identifies a higher risk, or for monogenic conditions, the addition of polygenic risk is unlikely to bring additional useful information. However, as the GWAS and PGS catalogues keep expanding their – for now limited – repertoire of traits and conditions, this approach could be especially meaningful for implementation in preventive care, where risk stratification targets a younger, usually healthier populations [69,70,71].

Second, the large sample size of the UK Biobank even after exclusion of individuals without follow-up, allows us to extrapolate the potential impact of these models for preventive lifestyle intervention at large scale. In the Netherlands, more than 16000 people enrolled themselves in a combined lifestyle intervention program between January 2019 and April 2020 alone. In a UKB population at least twice that, 9000 and 14000 fewer people would have been recommended lifestyle intervention by the decision rules compared to the clinical risk scores for T2D and hypertension. With a growing number of digital medical data and digital therapeutics platforms available to support clinicians and empower individuals to proactively act on their health, it is becoming easier to collected, process and analyse data from different sources such as the blood, body composition, and genetic markers evaluated in this study. When integrated with such platforms, the models developed in this study represent a viable, potentially less resource intensive framework for lifestyle interventions in preventive and primary care.

This study also presented some limitations. Firstly, the list of risk factors included is not exhaustive, due to both the high level of evidence required for inclusion in the model (most studies considered were meta-analyses) as well as the non-availability of other relevant variables in the UK Biobank data repository. Secondly, being a decision rules model, our proposed model does not provide individual risk predictions. While this increases the interpretability and applicability of the model (especially in a primary and preventive care setting), individuals within the same stratum may have different actual risk. Thirdly, we conducted the analysis with the assumption that all individuals classified as high risk who would have been recommended lifestyle intervention would not only have started it, but also achieved some degree of success. With a growing offer of consumer health and wellbeing programs, as well as employer-sponsored health programs, it is easier than ever before for individuals to preventively implement lifestyle changes [72]. However, many factors not accounted for here play a role in determining the actual effectiveness of these programs, so prospective validation in a study setting as well as in the market is required to assess the actual impact of these models on the effectiveness of preventive health interventions. Lastly, both the GWAS for the three PRS used in this study, as well as the UK Biobank cohort itself, are very ethnically homogeneous, with more than 90% of total participants being of white ethnicity and European descent. Therefore, the PRS results for UK Biobank participants of other ethnicities may be sub-optimal, and PRS and model validation will be required in cohorts with more diverse ethnical background.

In conclusion, in this prospective population-based cohort study of 60782 people, we developed and validated three risk stratification models for three prevalent chronic conditions. Adding other blood markers, physical measurements and genetic susceptibility scores to currently used clinical risk scoring tools resulted in moderate improvements in performance and in the identification of individuals likely to benefit from lifestyle intervention. When integrated with digital data or digital therapeutics platforms that enable the collection and analysis of these data, these algorithms can be used to support the successful adoption of lifestyle interventions in preventive and primary care.

## Data Availability

The data that support the findings of this study are available from the UK Biobank project site, subject to registration and application process. Further details can be found at https://www.ukbiobank.ac.uk.

## Acknowledgements

We thank participants in both the UK Biobank as well as all clinical, academic and administrative staff involved in data collection and storage.

## Author contributions

JCF: main contributor to all aspects of the manuscript. PF: main contributor to the data analysis and the methods and results sections of the manuscript. RG: original ideation of the manuscript, and significant contributions to the drafting of the manuscript. SK: original ideation. SM: significant contributor to the methods and discussion sections of the manuscript. TdK: co-supervisor in the clinical aspects of the manuscript, and significant contributor to the discussion and interpretation of the findings. SvD: significant contributor to the data analysis and the methods and results sections of the manuscript. Responsible for the data access request to UKB. BW: main supervisor in the clinical aspects of the manuscript, and significant contributions to the introduction and discussion. All authors gave input towards and approved the final manuscript.

## References

1) GBD 2017 Mortality Collaborators. Global, regional, and national age-specific mortality and life expectancy, 1950–2017: a systematic analysis for the Global Burden of Disease Study 2017. Lancet 2018; 392: 1684–735

2) WHO. Health systems performance assessment: debates, methods and empiricism. https://www.who.int/publications/2003/hspa/en/ (xaccessed October 30, 2020).

3) Global Burden of Disease Global, regional, and national incidence, prevalence, and years lived with disability for 354 diseases and injuries for 195 countries and territories, 1990–2017: a systematic analysis for the Global Burden of Disease Study 2017. Lancet. 2018; 392, 1789–1858.

4) Piepoli MF, et al. 2016 European Guidelines on cardiovascular disease prevention in clinical practice: The Sixth Joint Task Force of the European Society of Cardiology and Other Societies on Cardiovascular Disease Prevention in Clinical Practice (constituted by representatives of 10 societies and by invited experts) Developed with the special contribution of the European Association for Cardiovascular Prevention & Rehabilitation (EACPR). Eur Heart J. 2016; 37: 2315–2381.

5) Franklin BA, Cushman M. Recent Advances in Preventive Cardiology and Lifestyle Medicine. Circulation. 2011; 123:2274–2283.

6) Magkos F, Hjorth MF, Astrup A. Diet and exercise in the prevention and treatment of type 2 diabetes mellitus. Nature Reviews Endocrinology. 2020; 16: 545–555.

7) Wilson PW, D’Agostino RB, Levy D, Belanger AM, Silbershatz H, Kannel WB. Prediction of coronary heart disease using risk factor categories. Circulation. 1998; 97(18):1837–47.

8) Wilson PW, Meigs JB, Sullivan L, Fox CS, Nathan DM, D’Agostino RB. Prediction of incident diabetes mellitus in middle-aged adults: the Framingham Offspring Study. Archives of Internal Medicine. 2007;167(10):1068–74.

9) Parikh NI, Pencina MJ, Wang TJ, Benjamin EJ, Lanier KJ, Levy D, D’Agostino RB, Kannel WB, Vasan RS. A risk score for predicting near-term incidence of hypertension: the Framingham Heart Study. Annals of internal medicine. 2008;148(2):102–10

10) Damen JA, Pajouheshnia R, Heus P, Moons KGM, Reitsma JB, Scholten RJPM, et al. Performance of the Framingham risk models and pooled cohort equations for predicting 10-year risk of cardiovascular disease: a systematic review and meta-analysis. BMC Medicine. 2019; 17:109.

11) Damen JAAG, Hooft L, Schuit E, Debray TPA, Collins GS, Tzoulaki I, et al. Prediction models for cardiovascular disease risk in the general population: systematic review. BMJ. 2016; 16:353:i2416.

12) Mars NJ, Koskela JT, Ripatti P, Kiiskinen TTJ, Havulinna AS, Lindbohm JV, Ahola-Olli A, Kurki M, Karjalainen J, Palta P, et al. Polygenic and clinical risk scores and their impact on age at onset of cardiometabolic diseases and common cancers. Nat Med. 2020;26(4):549–557. doi: 10.1038/s41591-020-0800-0. Epub 2020 Apr 7.

13) Khera AV, Chaffin M, Aragam KG, Haas ME, Roselli C, Choi SH, et al. Genome-wide polygenic scores for common diseases identify individuals with risk equivalent to monogenic mutations. Nat Genet. 2018; 50: 1219–1224.

14) Chatterjee N, Shi J, Garcia-Closas M. Developing and evaluating polygenic risk prediction models for stratified disease prevention. Nat Rev Genet. 2016; 17: 392–406.

15) Rutten-Jacobs LC, Larsson SC, Malik R, Rannikmäe K, et al. Genetic risk, incident stroke, and the benefits of adhering to a healthy lifestyle: cohort study of 306 473 UK Biobank participants. BMJ. 2018 Oct 24;363:k4168. doi: 10.1136/bmj.k4168.

16) Said MA, Verweij N, van der Harst P. Associations of Combined Genetic and Lifestyle Risks With Incident Cardiovascular Disease and Diabetes in the UK Biobank Study. JAMA Cardiol. 2018 Aug 1;3(8):693–702. doi: 10.1001/jamacardio.2018.1717.

17) Lewis CM, Vassos E. Polygenic risk scores: from research tools to clinical instruments. Genome Med. 2020; 12: 44. https://doi.org/10.1186/s13073-020-00742-5

18) Whelton PK, Carey RM, Aronow WS, Casey Jr DE, Collins KJ, Himmelfarb CD, et al. 2017 ACC/AHA/AAPA/ABC/ACPM/AGS/APhA/ASH/ASPC/NMA/PCNA Guideline for the Prevention, Detection, Evaluation, and Management of High Blood Pressure in Adults: A Report of the American College of Cardiology/American Heart Association Task Force on Clinical Practice Guidelines. Hypertension. 2018;71:e13–e115. https://doi.org/10.1161/HYP.0000000000000065.

19) Nederlands Huisartsen Genootschap. Diabetes mellitus type 2, derde herziening 2018. [Internet]. Available from: https://richtlijnen.nhg.org/standaarden/diabetes-mellitus-type-2#volledige-tekst-literatuur [Accessed 15th July 2020].

20) Grundy SM, et al. 2018 AHA/ACC/AACVPR/AAPA/ABC/ACPM/ADA/AGS/APhA/ASPC/NLA/PCNA Guideline on the management of blood cholesterol: a report of the American College of Cardiology/American Heart Association Task Force on Clinical Practice Guidelines. J. Am. Coll. Cardiol. 73, 3168–3209 (2019).

21) Mach F, et al. 2019 ESC/EAS Guidelines for the management of dyslipidaemias: lipid modification to reduce cardiovascular risk. European Heart Journal. 2020; 41: 111–188.

22) Moher D, Liberati A, Tetzlaff J, Altman DG, PRISMA Group. Preferred reporting items for systematic reviews and meta-analyses: the PRISMA statement. Ann Intern Med 2009; 151:264-9, W64. doi:10.7326/0003-4819-151-4-200908180-00135

23) Emerging Risk Factors Collaboration, Di Angelantonio E, et al. Major lipids, apolipoproteins, and risk of vascular disease. JAMA. 2009; 302: 1993–2000.

24) Madsen CM, et al. Extreme High High-Density Lipoprotein Cholesterol Is Paradoxically Associated With High Mortality in Men and Women: Two Prospective Cohort Studies. Eur Heart J. 2017; 38 (32): 2478–2486.

25) Third Report of the National Cholesterol Education Program (NCEP) Expert Panel on Detection, Evaluation, and Treatment of High Blood Cholesterol in Adults (Adult Treatment Panel III) Final Report. Circulation. 2002; 106(25):3143.

26) Nordestgaard BG, Varbo A. Triglycerides and cardiovascular disease. Lancet. 2014; 384(9943):626–635.

27) Sarwar N, et al. Triglycerides and the Risk of Coronary Heart Disease: 10 158 Incident Cases Among 262 525 Participants in 29 Western Prospective Studies. Circulation. 2007; 115:450–458.

28) Ikezaki H, et al. Direct Versus Calculated LDL Cholesterol and C-Reactive Protein in Cardiovascular Disease Risk Assessment in the Framingham Offspring Study. Clin Chem. 2019; 65 (9): 1102–1114.

29) Penson PE, et al. Associations between very low concentrations of low-density lipoprotein cholesterol, high sensitivity C-reactive protein, and health outcomes in the Reasons for Geographical and Racial Differences in Stroke (REGARDS) study. European Heart Journal. 2018; 39: 3641–3653.

30) Arnett DK, et al. 2019 ACC/AHA Guideline on the Primary Prevention of Cardiovascular Disease: A Report of the American College of Cardiology/American Heart Association Task Force on Clinical Practice Guidelines. Circulation. 2019; 140 (11): e596–e646.

31) Richter B, Hemmingsen B, Metzendorf M-I, Takwoingi Y. Development of type 2 diabetes mellitus in people with intermediate hyperglycaemia. Cochrane Database Syst Rev. 2018; 10(10):CD012661. doi: 10.1002/14651858.CD012661.pub2.

32) Ren Y, Luo X, Wang C, Yin L, Pang C, Feng T. Prevalence of hypertriglyceridemic waist and association with risk of type 2 diabetes mellitus: a meta-analysis. Diabetes Metab Res Rev. 2016; 32(4):405–12. doi: 10.1002/dmrr.2725. Epub 2015 Nov 4.

33) Riaz H, Khan MS, Siddiqi. TJ, Usman MS, Shah N, Goyal A, et al. Association Between Obesity and Cardiovascular Outcomes: A Systematic Review and Meta-analysis of Mendelian Randomization Studies. JAMA Netw Open. 2018; 1(7):e183788. doi: 10.1001/jamanetworkopen.2018.3788.

34) Hashimoto Y, Hamaguchi M, Tanaka M, Obora A, Kojima T, Fukui M. Metabolically healthy obesity without fatty liver and risk of incident type 2 diabetes: A meta-analysis of prospective cohort studies. Obes Res Clin Pract. 2018; 12(1):4–15. doi: 10.1016/j.orcp.2017.12.003. Epub 2018 Jan 5.

35) Emdin CA, Anderson SG, Woodward M. Rahimi K. Usual Blood Pressure and Risk of New-Onset Diabetes: Evidence From 4.1 Million Adults and a Meta-Analysis of Prospective Studies. J Am Coll Cardiol. 2015; 66(14):1552–1562. doi: 10.1016/j.jacc.2015.07.059.

36) Reboussin DM, Allen NB, Griswold ME, Guallar E, Hong Y, Lackland DT, et al. Systematic Review for the 2017 ACC/AHA/AAPA/ABC/ACPM/AGS/APhA/ASH/ASPC/NMA/PCNA Guideline for the Prevention, Detection, Evaluation, and Management of High Blood Pressure in Adults: A Report of the American College of Cardiology/American Heart Association Task Force on Clinical Practice Guidelines. Circulation. 2018; 138(17):e595–e616. doi: 10.1161/CIR.0000000000000601.

37) Brunström M, Carlberg B. Association of Blood Pressure Lowering With Mortality and Cardiovascular Disease Across Blood Pressure Levels: A Systematic Review and Meta-analysis. JAMA Intern Med. 2018; 178(1):28–36. doi: 10.1001/jamainternmed.2017.6015.

38) Sundström J, Arima H, Jackson R, Turnbull F, Rahimi K, Chalmers J, et al. Effects of blood pressure reduction in mild hypertension: a systematic review and meta-analysis. Ann Intern Med. 2015; 162(3):184–91. doi: 10.7326/M14-0773.

39) Thomopoulos C, Parati G, Zanchetti A. Effects of blood pressure lowering on outcome incidence in hypertension: 2. Effects at different baseline and achieved blood pressure levels--overview and meta-analyses of randomized trials. J Hypertens. 2014; 32(12):2296–304. doi: 10.1097/HJH.0000000000000379

40) Hong Z, Wu T, Zhou S, Huang B, Wang J, Jin D, Geng D. Effects of anti-hypertensive treatment on major cardiovascular events in populations within prehypertensive levels: a systematic review and meta-analysis. J Hum Hypertens. 2018; 32(2):94–104. doi: 10.1038/s41371-017-0026-x. Epub 2018 Jan 9.

41) Deng G, Yin L, Liu W, Liu X, Xiang Q, Qian Z, et al. Associations of anthropometric adiposity indexes with hypertension risk: A systematic review and meta-analysis including PURE-China. Medicine (Baltimore). 2018; 97(48):e13262. doi: 10.1097/MD.0000000000013262.

42) Zhou Q, Shi Y, Li Y-Q, Ping Z, Wang C, Liu X, et al. Body mass index, abdominal fatness, and hypertension incidence: a dose-response meta-analysis of prospective studies. J Hum Hypertens. 2018; 32(5):321–333. doi: 10.1038/s41371-018-0046-1. Epub 2018 Mar 27.

43) Wei Y-C, George NI, Chang C-W, Hicks KA. Assessing Sex Differences in the Risk of Cardiovascular Disease and Mortality per Increment in Systolic Blood Pressure: A Systematic Review and Meta-Analysis of Follow-Up Studies in the United States. PLoS One. 2017; 12(1):e0170218. doi: 10.1371/journal.pone.0170218. eCollection 2017.

44) Auton A, Abecasis GR, Altshuler DM, Durbin RM, Bentley DR, Chakravarti A, et al. A global reference for human genetic variation. Nature. 2015; 526(7571):68–74. https://doi.org/10.1038/nature15393

45) Das S, Forer L, Schönherr S, Sidore C, Locke AE, Kwong A, et al. Next-generation genotype imputation service and methods. Nature Genetics. 2016; 48(10); 1284–1287. https://doi.org/10.1038/ng.3656

46) Hoffmann TJ, Ehret GB, Nandakumar P, Ranatunga D, Schaefer C, Kwok PY et al. Genome-wide association analyses using electronic health records identify new loci influencing blood pressure variation. Nature Genetics. 2017; 49(1):54–64. https://doi.org/10.1038/ng.3715

47) Nikpay M, Goel A, Won HH, Hall LM, Willenborg C, Kanoni S, et al. A comprehensive 1000 Genomes-based genome-wide association meta-analysis of coronary artery disease. Nature Genetics. 2015; 47(10):1121–1130. https://doi.org/10.1038/ng.3396

48) Scott RA, Scott LJ, Mägi R, Marullo L, Gaulton KJ, Kaakinen M, Pervjakova N, et al. An Expanded Genome-Wide Association Study of Type 2 Diabetes in Europeans. Diabetes. 2017; 66(11):2888–2902. https://doi.org/10.2337/db16-1253

49) DeLong ER, DeLong DM, Clarke-Pearson DL. Comparing the areas under two or more correlated receiver operating characteristic curves: a nonparametric approach. Biometrics. 1988; 44(3):837–845.

50) Lear SA, James PT, Ko GT, Kumanyika S. Appropriateness of waist circumference and waist-to-hip ratio cutoffs for different ethnic groups. Eur J Clin Nutr. 2010; 64(1):42–61. doi: 10.1038/ejcn.2009.70. Epub 2009 Aug 12.

51) Flack JM, Adekola B. Blood pressure and the new ACC/AHA hypertension guidelines. Trends Cardiovasc Med. 2020; 30(3):160–164. doi: 10.1016/j.tcm.2019.05.003

52) Zhang M, Zhang H, Wang C, Ren Y, Wang B, Zhang L, et al. Development and Validation of a Risk-Score Model for Type 2 Diabetes: A Cohort Study of a Rural Adult Chinese Population. PLoS ONE. 2016; 11(4):e0152054. https://doi.org/10.1371/journal.pone.0152054

53) Carroll SJ, Paquet C, Howard NJ, et al. Validation of continuous clinical indices of cardiometabolic risk in a cohort of Australian adults. BMC Cardiovasc Disord. 2014; 14:27. https://doi.org/10.1186/1471-2261-14-27

54) Udler MS, Kim J, von Grotthuss M, Bonàs-Guarch S, Cole JB, Chiou J, et al. (2018) Type 2 diabetes genetic loci informed by multi-trait associations point to disease mechanisms and subtypes: A soft clustering analysis. PLoS Med 15(9): e1002654. https://doi.org/10.1371/journal.pmed.1002654

55) Ahlqvist E, Storm P, Karajamaki A, Martinell M, Dorkhan M, Carlsson A, et al. Novel subgroups of adult-onset diabetes and their association with outcomes: a data-driven cluster analysis of six variables. Lancet Diabetes Endocrinol. 2018. doi: 10.1016/S2213-8587(18)30051-2.

56) Kivimäki M, Batty GD, Singh-Manoux A, Ferrie JE, Tabak AG, Jokela M, et al. Validating the Framingham Hypertension Risk Score: Results From the Whitehall II Study. Hypertension. 2009;54:496–501

57) Syllos DH, Calsavara VF, Bensenor IM, Lotufo PA. Validating the Framingham Hypertension Risk Score: A 4-year follow-up from the Brazilian Longitudinal Study of the Adult Health (ELSA-Brasil). J Clin Hypertens (Greenwich). 2020; 22(5):850–856. doi: 10.1111/jch.13855. Epub 2020 Apr 18

58) D’Agostino RB, Grundy S, Sullivan LM, Wilson P, CHD Risk Prediction Group. Validation of the Framingham Coronary Heart Disease Prediction Scores. Results of a Multiple Ethnic Groups Investigation. JAMA. 2001;286(2):180–187. doi:10.1001/jama.286.2.180

59) Koller MT, Leening MJG, Wolbers M, Steyerberg EW, Hunink M, Schoop R, et al. Development and Validation of a Coronary Risk Prediction Model for Older U.S. and European Persons in the Cardiovascular Health Study and the Rotterdam Study. Ann Intern Med. 2012; 157(6):389–397. doi: 10.7326/0003-4819-157-6-201209180-00002.

60) Lean ME, Leslie WS, Barnes AC, Brosnahan N, Thom G, McCombie L, et al. Primary care-led weight management for remission of type 2 diabetes (DiRECT): an open-label, cluster-randomised trial. Lancet. 2018; 391(10120):541–551. doi: 10.1016/S0140-6736(17)33102-1. Epub 2017 Dec 5.

61) Kim SE, Castro Sweet CM, Cho E, Tsai J, Cousineau MR. Evaluation of a Digital Diabetes Prevention Program Adapted for Low-Income Patients, 2016-2018. Prev Chronic Dis. 2019;16:E155. doi: 10.5888/pcd16.190156.

62) Athinarayanan SJ, Adams RN, Hallberg SJ, McKenzie AL, Bhanpuri NH, Campbell WW, Volek JS, Phinney SD and McCarter JP. Long-Term Effects of a Novel Continuous Remote Care Intervention cluding Nutritional Ketosis for the Management of Type 2 Diabetes: A 2-Year Non-randomized Clinical Trial. 2019. Front Endocrinol; 10:348.

63) McCarthy MI. Painting a new picture of personalised medicine for diabetes. Diabetologia. 2017; 60(5):793-799. DOI: 10.1007/s00125-017-4210-x.

64) Abraham G, Malik R, Yonova-Doing E, et al. Genomic risk score offers predictive performance comparable to clinical risk factors for ischaemic stroke. Nat Commun. 2019; 10: 5819

65) Müller-Riemenschneider F, Holmberg C, Rieckmann N, et al. Barriers to Routine Risk-Score Use for Healthy Primary Care Patients: Survey and Qualitative Study. Arch Intern Med. 2010; 170(8):719– 724. doi:10.1001/archinternmed.2010.66.

66) Kappen TH, Van Loon K, Kappen MA, et al. Barriers and facilitators perceived by physicians when using prediction models in practice. J Clin Epidemiol. 2016. 70: 136–145.

67) Rosselo X, Dorresteijn JAN, Janssen A, et al. Risk prediction tools in cardiovascular disease prevention. European Journal of Preventive Cardiology. 2019; 26(14):1534–1544.

68) Lloyd-Jones DM, Leip EP, Larson MG, D’Agostino RB, Beiser A, Wilson PWF, et al. Prediction of Lifetime Risk for Cardiovascular Disease by Risk Factor Burden at 50 Years of Age. Circulation. 2006; 113:791–798.

69) MacArthur J, Bowler E, Cerezo M, Gil L, Hall P, Hastings E, et al. The new NHGRI-EBI Catalog of published genome-wide association studies (GWAS Catalog). Nucleic Acids Res. 2017; 45:D896–D901

70) Lambert SA, Gil L, Jupp S, et al. The Polygenic Score Catalog as an open database for reproducibility and systematic evaluation. Nat Genet. 2021. https://doi.org/10.1038/s41588-021-00783-5

71) Levin MG, Rader DJ. Polygenic Risk Scores and Coronary Artery Disease: Ready for Prime Time? Circulation. 2020;141:637–640

72) Cohen AB, Mathews SC, Dorsey ER, Bates DW, Safavi K. Direct-to-consumer digital health. Lancet Digit Health. 2020; 2(4):e163–e165. doi: 10.1016/S2589-7500(20)30057-1. Epub 2020

