## Supplementary material for "Development and validation of decision rules models to stratify coronary artery disease, diabetes, and hypertension risk in preventive care: cohort study of returning UK Biobank participants"

### Appendix S1. TRIPOD Checklist.

| Section/Topic | ı | Checklist Item | Page |
|---|---|---|---|
| **Title and abstract** | | | |
| Title | 1 | Identify the study as developing and/or validating a multivariable prediction model, the target population, and the outcome to be predicted. | 1 |
| Abstract | 2 | Provide a summary of objectives, study design, setting, participants, sample size, predictors, outcome, statistical analysis, results, and conclusions. | 3 |
| **Introduction** | | | |
| Background and objectives | 3a | Explain the medical context (including whether diagnostic or prognostic) and rationale for developing or validating the multivariable prediction model, including references to existing models. | 5 |
| | 3b | Specify the objectives, including whether the study describes the development or validation of the model or both. | 6 |
| **Methods** | | | |
| Source of data | 4a | Describe the study design or source of data (e.g., randomized trial, cohort, or registry data), separately for the development and validation data sets, if applicable. | 7 |
| | 4b | Specify the key study dates, including start of accrual; end of accrual; and, if applicable, end of follow-up. | 7 |
| Participants | 5a | Specify key elements of the study setting (e.g., primary care, secondary care, general population) including number and location of centres. | 7 |
| | 5b | Describe eligibility criteria for participants. | 7 |
| | 5c | Give details of treatments received, if relevant. | N/A |
| Outcome | 6a | Clearly define the outcome that is predicted by the prediction model, including how and when assessed. | 10 |
| | 6b | Report any actions to blind assessment of the outcome to be predicted. | N/A |
| Predictors | 7a | Clearly define all predictors used in developing or validating the multivariable prediction model, including how and when they were measured. | 8,9,10 |
| | 7b | Report any actions to blind assessment of predictors for the outcome and other predictors. | N/A |
| Sample size | 8 | Explain how the study size was arrived at. | N/A |
| Missing data | 9 | Describe how missing data were handled (e.g., complete-case analysis, single imputation, multiple imputation) with details of any imputation method. | N/A |
| Statistical analysis methods | 10a | Describe how predictors were handled in the analyses. | 10 |
| | 10b | Specify type of model, all model-building procedures (including any predictor selection), and method for internal validation. | 8,9,10 |
| | 10d | Specify all measures used to assess model performance and, if relevant, to compare multiple models. | 10,11 |
| Risk groups | 11 | Provide details on how risk groups were created, if done. | |
| **Results** | | | |

| | | | |
|---|---|---|---|
| Participants | 13a | Describe the flow of participants through the study, including the number of participants with and without the outcome and, if applicable, a summary of the follow-up time. A diagram may be helpful. | 12 |
| | 13b | Describe the characteristics of the participants (basic demographics, clinical features, available predictors), including the number of participants with missing data for predictors and outcome. | 12 |
| Model development | 14a | Specify the number of participants and outcome events in each analysis. | 12 |
| | 14b | If done, report the unadjusted association between each candidate predictor and outcome. | 12,13 |
| Model specification | 15a | Present the full prediction model to allow predictions for individuals (i.e., all regression coefficients, and model intercept or baseline survival at a given time point). | N/A |
| | 15b | Explain how to the use the prediction model. | 8, 9, Append ix S2 |
| Model performance | 16 | Report performance measures (with CIs) for the prediction model. | 12,13 |
| **Discussion** | | | |
| Limitations | 18 | Discuss any limitations of the study (such as nonrepresentative sample, few events per predictor, missing data). | 18 |
| Interpretation | 19b | Give an overall interpretation of the results, considering objectives, limitations, and results from similar studies, and other relevant evidence. | 15,16, 17 |
| Implications | 20 | Discuss the potential clinical use of the model and implications for future research. | 16,17 |
| **Other information** | | | |
| Supplementary information | 21 | Provide information about the availability of supplementary resources, such as study protocol, Web calculator, and data sets. | 20 |
| Funding | 22 | Give the source of funding and the role of the funders for the present study. | 20 |

**Appendix S2.**

| **Coronary artery disease risk** | | |
|---|---|---|
| Risk stratum | Variables | Values |
| High risk | Total cholesterol | > 8 mmol/L |
| | LDL cholesterol | > 4.9 mmol/L |
| | Systolic blood pressure | > 180 mmHg |
| | Clinical risk score<br>Biomarkers other than total cholesterol or LDL | High<br>High |
| | Clinical risk score<br>PRS | High<br>$9^{th}$ or $10^{th}$ decile |
| No elevated risk | Biomarkers<br>Family history<br>Clinical risk score<br>PRS | All withing normal range<br>Negative<br>Low risk<br>< $8^{th}$ decile |
| Elevated risk | All other combinations where at least one risk factor is outside normal range | |

| **Diabetes risk** | | |
|---|---|---|
| Risk stratum | Variables | Values |
| High risk | HbA1c<br>Fasting glucose | > 6.5%<br>< 6.1 mmol/L |
| | Fasting glucose | > 6.1 mmol/L |
| | HbA1c<br>Weight | 5.5% - 6.4%<br>Overweight/obese |
| | Fasting glucose<br>Weight | 5.6 mmol/L - 6.1mmol/L<br>Overweight/Obese |

| | | |
|---|---|---|
| | HbA1c<br>PRS | 5.5% - 6.4%<br>$9^{th}$ or $10^{th}$ decile |
| | Fasting glucose<br>PRS | 5.6 mmol/L - 6.1mmol/L<br>$9^{th}$ or $10^{th}$ decile |
| | Clinical risk score | High |
| | Clinical risk score<br>Age<br>Triglycerides<br>HDL | Elevated<br>> 45<br>> 2.8 mmol/L<br>< 0.9 mmol/L |
| No elevated risk | Fasting glucose<br>HbA1c<br>Clinical risk score<br>PRS | < 5.6 mmol/L<br>< 5.5%<br>Low risk<br>< $8^{th}$ decile |
| Elevated risk | All other combinations where at least one risk factor is outside normal range | |

| **Hypertension risk** | | |
|---|---|---|
| Risk stratum | Variables | Values |
| High risk | Systolic blood pressure | 130-140 mmHg |
| | Diastolic blood pressure | 80-90 mmHg |
| | Clinical risk score | High |
| | Clinical risk score<br>PRS | Elevated<br>$9^{th}$ or $10^{th}$ decile |
| No elevated risk | Systolic blood pressure<br>Diastolic blood pressure<br>Clinical risk score<br>PRS | < 130 mmHg<br>< 80 mmHg<br>Low risk<br>< $8^{th}$ decile |

| | |
|---|---|
| Elevated risk | All other combinations where at least one risk factor is outside normal range |

**Appendix S3. Supplemental methods for polygenic risk score calculation**

Polygenic risk scores were created following an additive model for CAD, diabetes and hypertension. To calculate the PGS, the LDpred tool was used following the typical workflow of coordinating the required files, adjustment of SNP weights based on LD and the calculation of the PGS (Vilhjálmsson et al., 2015). The genotyping data and data containing the tested phenotypes outcomes were downloaded from the UKB. All variants with an imputation $R^2 < 0.4$ were removed based on the minimac3 reported $R^2$ readily available in the downloaded genotyping files, using plink (Purcell et al). In total, 3 PGS were calculated for CAD, diabetes, hypertension using summary statistics files referenced in the manuscript. The latter study is a GWAS on blood pressure, which we used to generate PGSs for hypertension. The respective summary statistics files were downloaded and, where necessary, reformatted to be consistent with the format required by LDpred. All variants with a GWAS significance p-value below 0.01 were selected (table S4). SNP ids where converted based on their chromosome, position, ref and alt alleles, to ensure no SNPs were lost due to inconsistent SNP naming between the files.

The Phase 3 genome files were used as an LD reference panel. The overlapping genotypes in the diabetes summary statistics file, the LD reference panel and the UKB genotyping files were extracted and coordinated using the LDpred coordinate option. This was followed by a reweighing step for the SNPs based on their LD. LDpred-inf option of LDpred version 1.0.11 was used to calculate the PGS scores where the causal fraction -f was set to 1, the LD region -ldf 1000 as suggested in their guidelines. The calculations of the PGS scores described above were conducted on a per chromosome basis. Afterward, the scores for all chromosomes were summed resulting in 1 PGS file containing a PGS for each individual. Next, we calculated the odds ratios between the 10% individuals with the highest risk against the rest of the population. We have conducted this analysis with and without adjustment for genotyping array, first 4 principal components, age and sex. We determined the risk increase that can be derived from solely genetic data and compared this to the risk increase predicted based on genetic data, age and sex (table S5). These results are in line with those reported by others (fig S4).

Table S1. **Search strategy and results.**

| Module | Search term | Time span | Articles found | Search date |
|---|---|---|---|---|
| Diabetes | ((diabetes [TIAB] AND type 2 [TIAB]) OR impaired glucose [TIAB] OR high blood glucose [TIAB]) AND (biomarker* [TIAB] OR risk [TIAB] OR Hazard [TIAB] OR Odds [TIAB]) AND Meta-analysis [pt] AND 2014:2019 [dp] | 2014-Dec 2019 | 1103 | 24-Dec 2019 |
| Hypertension | (hypertension [TIAB] OR high blood pressure [TIAB]) AND (biomarker* [TIAB] OR risk [TIAB] OR Hazard [TIAB] OR Odds [TIAB]) AND Meta-analysis [pt] AND 2014:2019 [dp] | 2014-Dec 2019 | 1225 | 24-Dec 2019 |
| Coronary artery disease | (atherosclerosis [TIAB] OR plaque build-up [TIAB] OR plaque buildup [TIAB] OR coronary artery disease [TIAB] OR coronary heart disease [TIAB] OR coronary event* [TIAB] OR cardiovascular event* [TIAB]) AND (biomarker* [TIAB] OR risk [TIAB] OR Hazard [TIAB] OR Odds [TIAB]) AND Meta-analysis [pt] AND 2014:2019 [dp] | 2014-Dec 2019 | 1747 | 30-Dec-2019 |

Figure S1. **PRISMA flow for the search for coronary artery disease.**

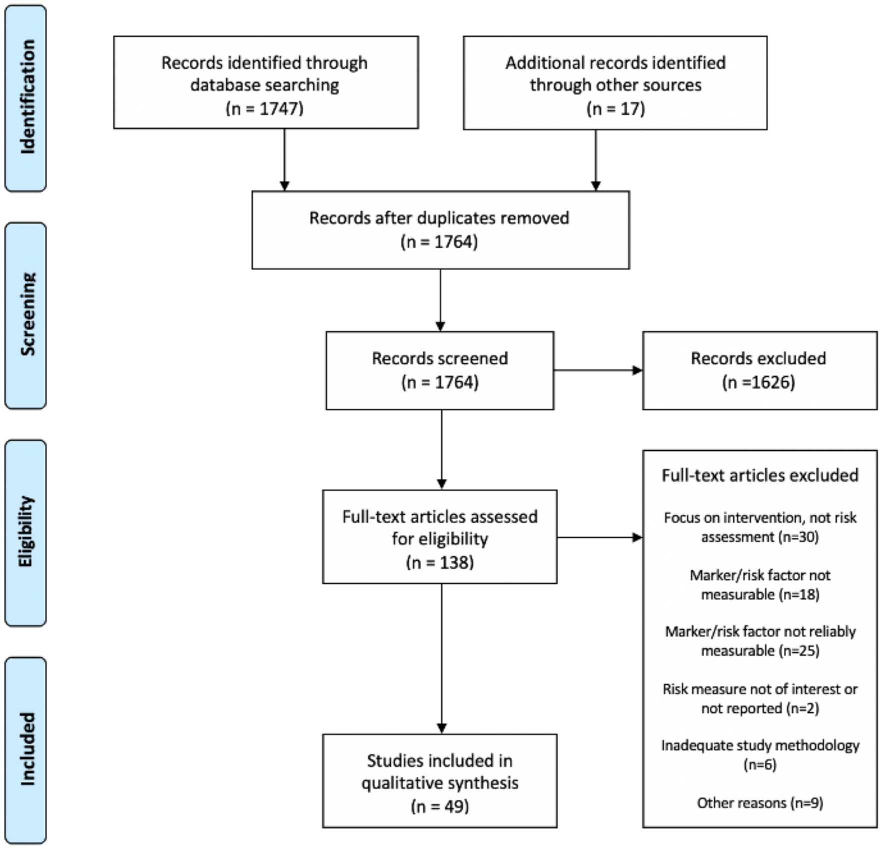

The search resulted in 1747 unique citations, with an additional 17 being identified manually. All of the abstracts were reviewed to see if they met the selection criteria. Then the full text of the preliminary selected articles was reviewed using the selection criteria, which resulted in a final identification of 138 publications. The full texts of 138 preliminarily selected articles were reviewed, resulting in a final identification of 49 relevant publications. A bibliography of these 49 publications is included below. The relevant national guideline identified as relevant for this module was the Dutch Society of General Practitioners Guideline for cardiovascular risk management (NHG Cardiovasculair risicomanagement richtlijn). This national guideline was based on the European Society of Cardiology "Guidelines for the management of dyslipidaemias: lipid modification to reduce cardiovascular risk", which were also deemed relevant.

Figure S2. **PRISMA flow for the search for hypertension.**

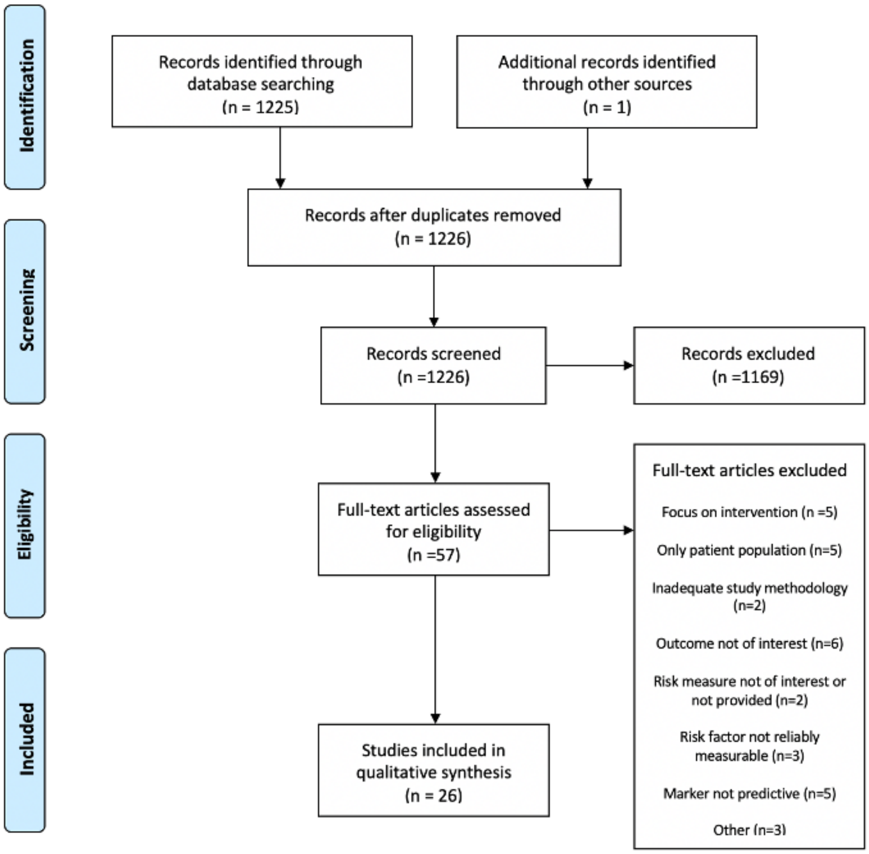

The search resulted in 1225 unique citations, with one additional citation being identified manually. All abstracts were reviewed to see if they met the selection criteria. Then the full text of the preliminary selected articles was reviewed using the selection criteria, which resulted in a final identification of 57 publications. The full texts of 57 preliminarily selected articles were reviewed using the selection criteria outlined in Appendix A and resulted in a final identification of 26 relevant publications. A bibliography of these 26 publications is included below.

Figure S3. **PRISMA flow for the search for diabetes**.

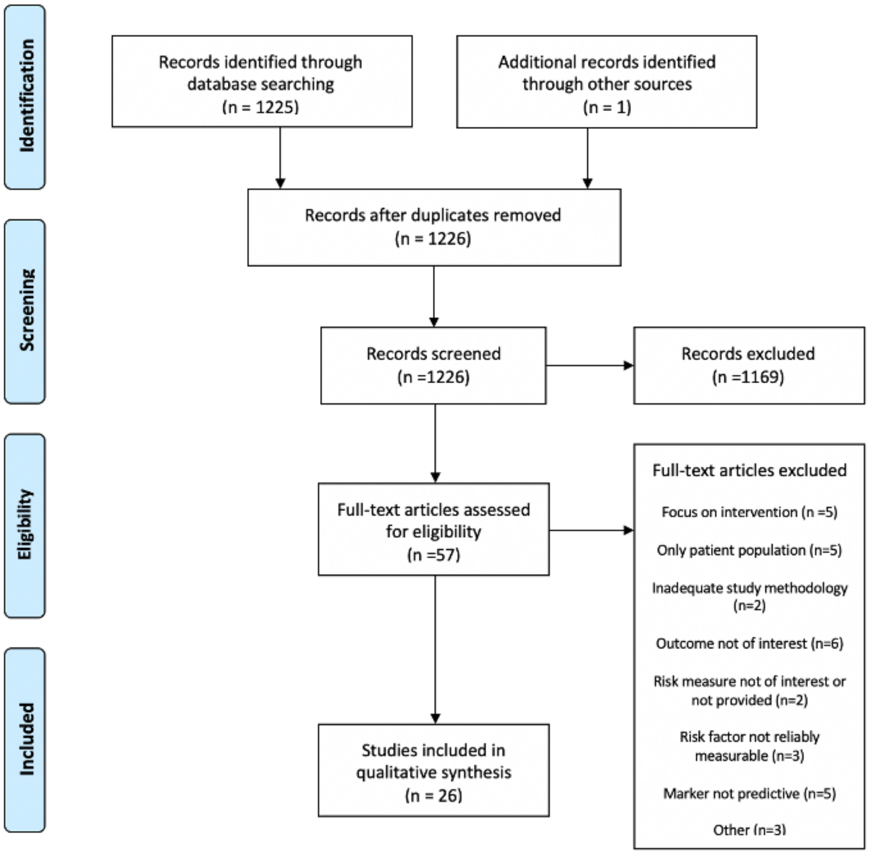

The search resulted in 1103 unique citations, with an additional 3 identified manually. All abstracts were reviewed to see if they met the selection criteria. Then the full text of the preliminary selected articles was reviewed using the selection criteria, which resulted in a final identification of 86 publications. The full texts of 86 preliminarily selected articles were reviewed, resulting in a final identification of 42 relevant publications. The Dutch General Practitioners' association workgroup guideline on type 2 diabetes defines clinical and subclinical thresholds for thresholds for fasting plasma glucose, in accordance with the World Health Organisation/International Diabetes Federation 2006 guideline. These cut-offs of 6.1 mmol/L and 7 mmol/L were taken in the module as "high risk" and "clinical thresholds" respectively.

Table S2. **UKB columns used to calculate disease incidence.**

| **Disease** | **Column(s)** | **Data-Field(s)** | **Value** |
|---|---|---|---|
| Diabetes | Diabetes diagnosed by doctor | 2443 | yes |
| | Non-cancer illness code, self-reported | 20002 | type 2 diabetes |
| | ICD10 | 41270, 41202, 41204, 40001, 40002 | E11 and related sub-codes |
| Hypertension | Vascular/heart problems diagnosed by doctor | 6150 | high blood pressure |
| | Non-cancer illness code, self-reported | 20002 | hypertension |
| | Medication for cholesterol, blood pressure or diabetes | 6177 | blood pressure medication |
| | ICD10 | 41270, 41202, 41204, 40001, 40002 | E10 |
| CAD | Vascular/heart problems diagnosed by doctor | 6150 | heart attack |
| | Non-cancer illness code, self-reported | 20002 | heart attack |
| | Operation code | 20004 | PTCA, CABG, triple heart bypass |
| | ICD9 | 41271, 41203, 41205 | 410-412 |

| | | | |
|---|---|---|---|
| | ICD10 | 41270, 41202, 41204, 40001, 40002 | I21-24, I25.2 |
| | OPCS4 | 41272, 41200, 41210 | K40-46, K50.1, K75 |

Table S3. **UKB age of diagnosis columns used to calculate disease incidence.**

| **Disease** | **Column(s)** | **Data-Field(s)** |
|---|---|---|
| Diabetes | Age diabetes diagnosed | 2976 |
| | Interpolated Year when non-cancer illness first diagnosed | 20008 |
| | ICD10 date of diagnosis | 41280, 41262, |
| Hypertension | Age high blood pressure diagnosed | 2966 |
| | Interpolated Year when non-cancer illness fiirst diagnosed | 20008 |
| | ICD10 date of diagnosis | 41280, 41262 |
| CAD | Age heart attack diagnosed | 3894 |
| | Interpolated Year when non-cancer illness first diagnosed | 20008 |
| | Interpolated Year when operation took place | 20010 |
| | ICD9 date of diagnosis | 41281, 41263 |
| | ICD10 date of diagnosis | 41280, 41262, |
| | OPCS4 date of diagnosis | 41282, 41260 |

Table S4. **Summary statistics data.** The column denotes the phenotype the PGS was used for. Followed by the number of cases and controls used in the GWAS study. The last three columns describe the total number of SNPs in the summary statistic file, those with a p-value <0.01 and those overlapping with the ones present in the LD reference panel and the UKB genotyping chip.

| **Phenotype** | **Cases** | **Controls** | **SNPs** | **p<0.01** | **Overlapping** |
|---|---|---|---|---|---|
| **Diabetes** | 26676 | 132532 | 12056346 | 199120 | 159290 |
| **CAD** | 60801 | 123504 | 9455805 | 139885 | 105325 |
| **Hypertension** | 80792 | 99785 | 28276170 | 400016 | 243528 |

CAD = coronary artery disease, SNP = single nucleotide polymorphism.

Table S5. **Risk for the individuals in the highest risk decile for each genetic risk score, compared to rest of population for the complete UKB population (n=442 687)**. Second and third column show hazard ratios calculated based on a logistic regression model adjusted for the respective variables. In all cases the difference with the remainder of the population was statistically significant (p-value < E-100).

| | **Unadjusted PRS** | **PRS adjusted for 4 PCs and array type** | **PRS adjusted for 4 PCs, array type, sex and age** | **Age and sex** |
|---|---|---|---|---|
| **CAD** | 1.99 (1.93-2.04) | 2.06 (2-2.11) | 4.77 (4.68-4.86) | 3.74 (3.66-3.82) |
| **T2D** | 1.93 (1.88-1.98) | 2.05 (2-2.09) | 2.90 (2.84-2.96) | 2.22 (2.17-2.28) |
| **Hypertension** | 1.37 (1.35-1.39) | 1.38 (1.37-1.4) | 1.85 (1.83-1.87) | 1.66 (1.64-1.68) |

Figure S4. **Prevalence of disease of individuals classed into different risked strata based on their genetic susceptibility scores.**

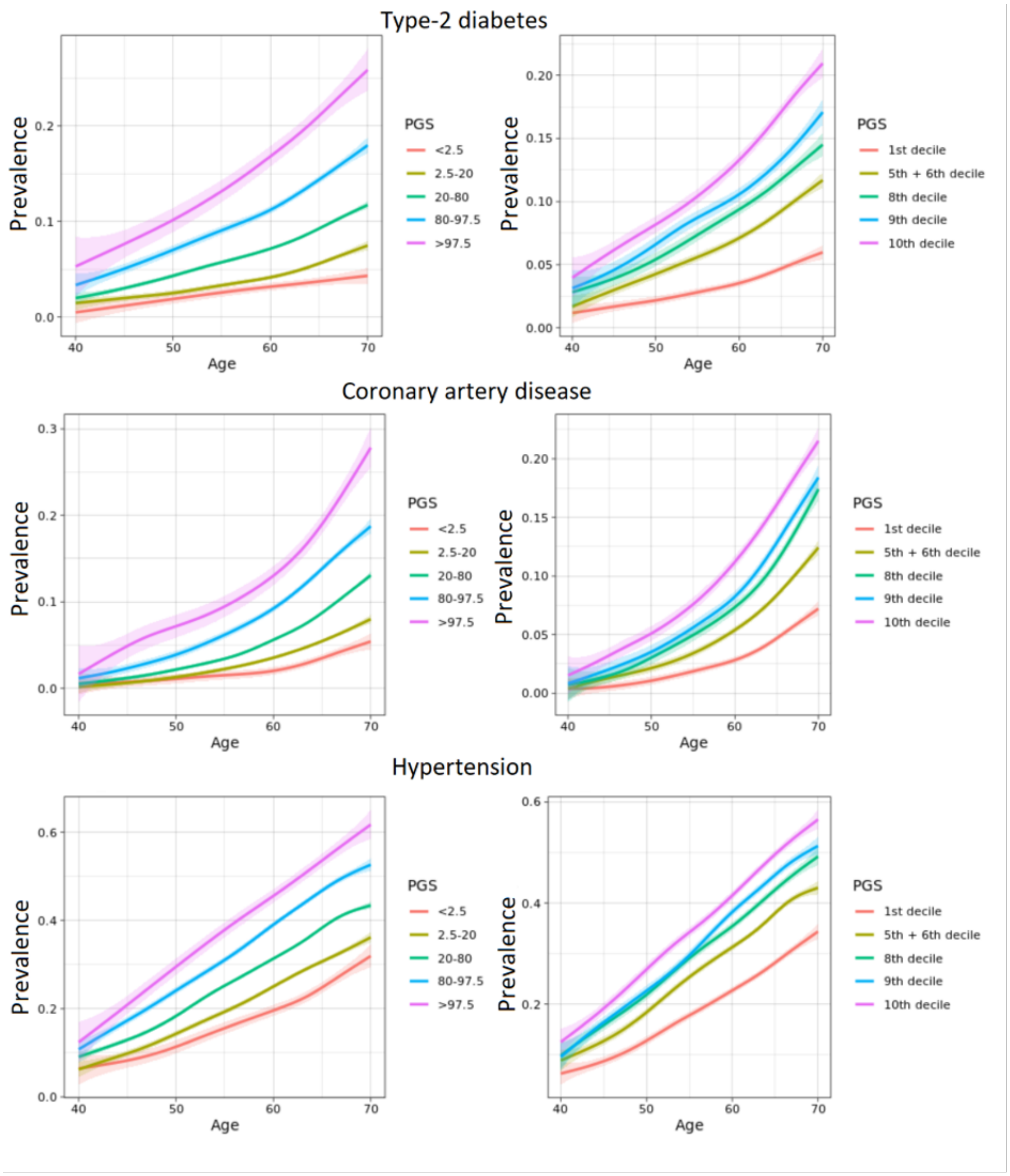

Left side shows risk strata as defined in Mars et al. Scores were corrected for first 4 principal components and array type. Only individuals with follow-up data are included (table 3). The AUROCs reported in table 2 pertain to the comparison between the 1-7th deciles and the 10th decile (table 2).
